# Characteristics and costs of electric scooter injuries in Helsinki: a retrospective cohort study

**DOI:** 10.1101/2022.06.14.22276168

**Authors:** Henri Vasara, Linda Toppari, Veli-Pekka Harjola, Kaisa Virtanen, Maaret Castrén, Arja Kobylin

## Abstract

**Background and purpose:** The incidence of electric scooter (e-scooter) injuries has increased drastically in numerous countries after widespread availability of shared e-scooters. Most publications on e-scooter injuries focus on a specific category of injuries. The economic impact on society from a broader perspective has not been studied.

**Patients and methods:** We performed a retrospective cohort study including all e-scooter-related injuries presented in the three adult emergency departments in Helsinki in 2021. We collected the patient data from the university hospital information system. Injury severity was evaluated based on the abbreviated injury score. Cost of the hospital treatment was analyzed based on our hospital district’s service price listing. In addition, we recorded the total amount of sick leave days and estimated their economic impact.

**Results:** In total, 446 e-scooter injuries were identified and taken into the analysis (434 affecting riders and 12 non-riders). The median age of the patients was 26 (IQR 22-33) and 59 % were male. 257 (58 %) of the of the injuries were minor, whereas 155 (35 %) were moderate, 30 (7 %) serious, 3 (0.7 %) severe, and one (0.2 %) critical. Furthermore, 220 (49 %) of the patients sustained head injuries. A major spike in accident incidence was seen during weekend (Friday to Sunday) nights, which was accompanied by a proportional increase in patients with alcohol intoxication. Including both the costs of the hospital care and absence from work, the approximated total cost of e-scooter injuries was 1.7 million euros, with a median cost of a single accident being 1148 euros (IQR 399 – 4263 €).

**Interpretation:** Comprehensive preventive measures must be conducted to decrease the incidence of e-scooter injuries. The usage of helmets should be strongly encouraged in order to prevent severe head injuries. The night-time bans during weekends and speed-limits on e-scooters appear to be justifiable.

## Background

Electric scooters (e-scooters) have become a widespread means of transport globally, especially in large metropolitan cities. The arrival of shared e-scooter rental companies has contributed largely to the popularity of this emerging mode of transportation. (Haworth et al., 2021). Unfortunately, as a by-product, e-scooter-related accidents have increased drastically in numerous countries (Badeau et al., 2019; Coelho et al., 2021; Farley et al., 2020; Namiri et al., 2020; Shichman et al., 2022).

The incidence of e-scooter-related accidents has been reported as 60 injuries per 100 000 rides (Bekhit et al., 2020). In most countries, the overall incidence has increased rapidly after the introduction of shared e-scooter companies (Farley et al., 2020; Shichman et al., 2022). Compared to cycling, the risk for injuries is approximately 3.8 times higher (Cicchino et al., 2021). The most typical injuries consist of head injuries, fractures of the extremities, and superficial wounds. However, more severe injuries requiring intensive care or even fatality have been reported. (Moftakhar et al., 2021; Shichman et al., 2021, 2022; Siow et al., 2020; Trivedi et al., 2019)

There has been a rising number of publications on e-scooter injuries in recent years. However, most studies focus solely on a single category of injuries (Factor et al., 2021; Suominen et al., 2022) or cases from a single hospital (Shichman et al., 2021). To our knowledge, the incidence of accidents in proportion e-scooter usage has been estimated only by Bekhit et al. (Bekhit et al., 2020).

In the current literature, there are only a few cost estimates for e-scooter injuries, and they vary greatly depending on the country (Bekhit et al., 2020; Lavoie-Gagne, Siow, Harkin, Flores, Politzer, et al., 2021). However, these numbers focus solely on the cost of the hospital treatment. Economical impact on society from a broader perspective has not been studied.

The objective of this study was to estimate the incidence of e-scooter injuries requiring hospital treatment and to describe the injury patterns and severity. In addition, we aimed to estimate the costs of e-scooter injuries to the healthcare system and society.

## Patients and methods

### Setting

We performed a retrospective cohort study assessing all e-scooter-related injuries in the Helsinki between January 2021 and December 2021. We analyzed all patients enrolled in the adult emergency departments (ED): one level 1 trauma center (responsible for high-level trauma care for the whole Hospital district in southern Finland), and two level 4 trauma centers (responsible for mild and moderate-level trauma patients).

The population base of Helsinki was 656 920 residents on 31.12.2020, with the mean age being 41 years and 53 % being female (Jaakkola et al., 2021). During the study period, five shared e-scooter rental companies with a total fleet of approximately 4700 e-scooters were in operation. The e-scooters were rented via a mobile application for users over 18 years old. In Finland, the maximum speed of e-scooters is 25km/h by law. Furthermore, the use of helmets is strongly encouraged but are not officially controlled.

During the study period the city of Helsinki and the e-scooter rental companies constituted several restrictions for rental e-scooter usage. First, on 7^th^ of July, the maximum speed limit was lowered to 15 km/h on several inner-city areas. Second, on 3^rd^ of September, the use of rental e-scooters was prohibited on Friday and Saturday nights between 00.00 and 05.00. In addition, the top speed was lowered to 20 km/h during daytime and to 15 km/h between 00.00 and 05.00, except on Fridays and Sundays.

### Data inquiry

We performed an initial data inquiry to detect e-scooter-related injuries using a keyword search from the patient datapool. Four different e-scooter-related words, including their inflected forms, were used. Next, the authors HV and LT examined all the detected patient records. Consequently, only the cases where the e-scooter involvement was definite were included. All remaining patients over 16 years old were included in the study. Exclusive inclusion and exclusion criteria are presented in figure 1.

**Figure 1:**
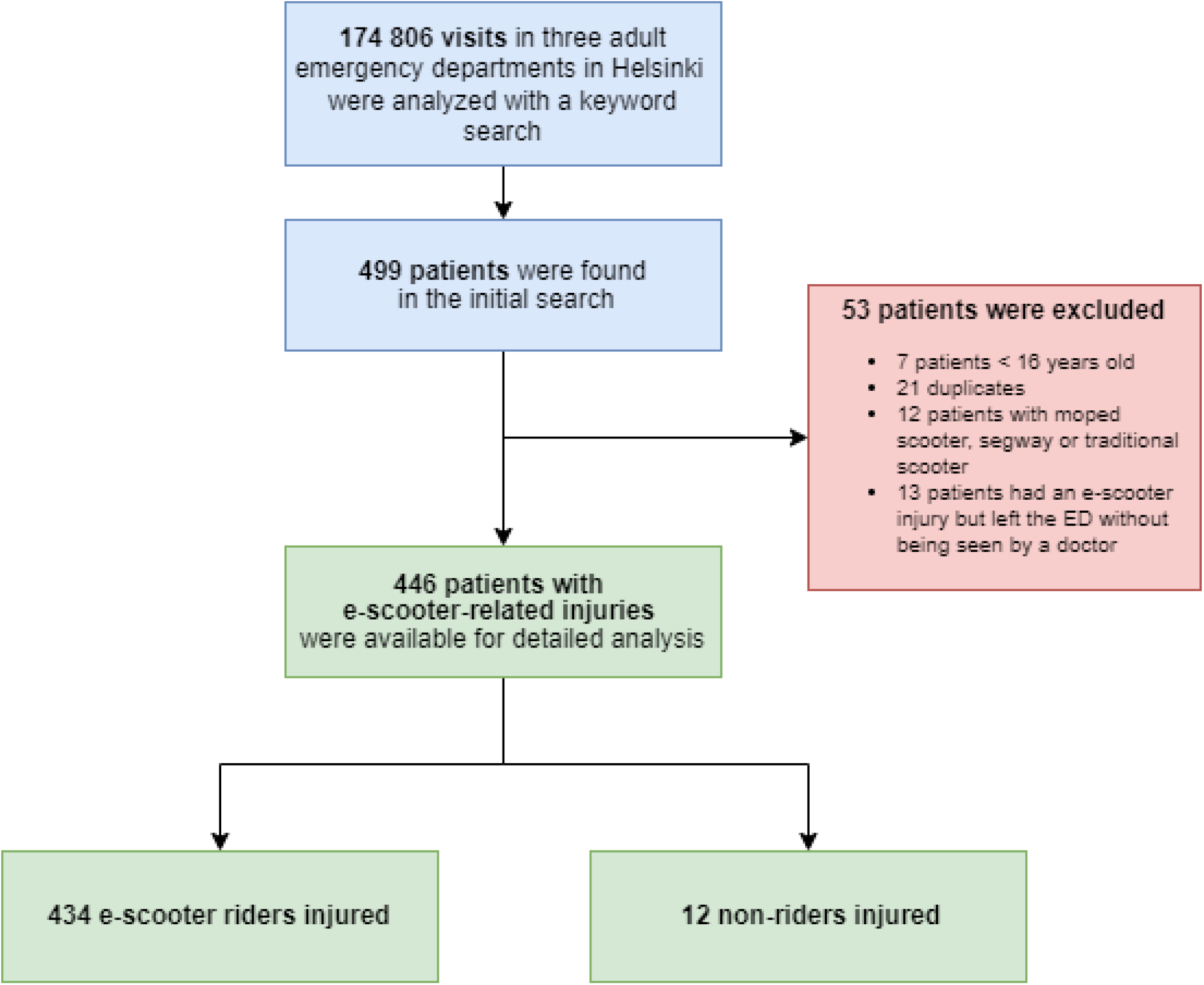
Flow chart of the Inclusion and exclusion criteria.

### Patient demographics

We collected the demographic factors from our hospital information system and ambulance records at the time of the ED visit. The collected variables were age, sex, time of the injury, injury mechanism, the person affected in the injury, helmet usage, and alcohol intoxication status. If the exact time of injury was not available, we set the injury time as the time when the patient registered at the ED. The breath alcohol level was recorded if measured at the ED or ambulance. In addition, alcohol intoxication was assessed as a binominal value considering the clinical assessment of the ED doctors if the breath alcohol level was not measured.

### Injury patterns and severity

We collected all sustained injuries based on the diagnoses from the e-scooter hospital episode according to the International Classification of Diagnoses-10 (ICD-10). In case of missing diagnoses, we added the injuries if they were described with enough precision in the patient records. The injuries are presented based on their anatomical location and severity.

Every patient’s most severe injury was graded based on Abbreviated Injury Score (AIS) from the Abbreviated Injury Scale (Association for the Advancement of Automotive Medicine, 2015) to minor, moderate, serious, severe, or critical. Superficial wounds, muscle or ligament sprains, mild contusions, or equivalent were graded as minor. Fractures of distal extremities, head concussions, and equivalent were graded as moderate injuries. Open fractures, intracranial hemorrhages requiring no surgical interventions, comminuted fractures, or fractures in load-bearing bones were graded as serious injuries. Furthermore, intracranial hemorrhages requiring craniotomy and patients requiring multiple days of intensive care were graded as severe or critical.

To estimate the total effect of all injuries on the patient, we calculated the Injury Severity Score (ISS) (Osler et al., 1997). Accordingly, patients with ISS 1-8 were graded as minor, ISS 8-13 were graded as moderate, and ISS > 13 were graded as major trauma patients, respectively.

### Follow-up

After the initial ED visit, we followed up the patients from our hospital information system for a minimum of 2 months or to the end of the e-scooter-related hospital episode if the treatment continued beyond 2 months. If the patient required hospital admissions, we recorded the length and type of the hospital admission. In addition, all scheduled additional injury-related hospital visits and surgeries were recorded and analyzed. We recorded the total amount of radiographs (native radiographs, computed tomography scans, and magnetic resonance imaging), laboratory tests, and the amount of sickness absence days the physician dictated for each patient during all hospital visits.

### Cost analysis

The estimation of costs for the whole treatment (ED visits, inpatient care, outpatient visits, surgical procedures, radiological imaging, and laboratory tests) was based on our hospital district’s service price listing for 2021 (Helsinki University Hospital, 2021). In our hospital district, the service price listing describes the costs that are billed from the patients’ home city after their care. A price was sought for all events during the hospital episode. If the price of an event was unequivocal, the lowest suitable value was used.

According to the Finnish social system, the pay for short-term absence (≤ 10 working days) is covered by the employer, and the Social Insurance Institution of Finland covers long-term absence (> 10 days). To further estimate the costs of sick leave for the work providers and society, we used the Finnish Ministry of Social Affairs and Health evaluation in 2014. Accordingly, we used an inflation corrected price of 203,91 €/day for short-term sickness absence and 158.83€/day for long-term sickness absence. (Rissanen & Kaseva, 2014; Statistics Finland, 2021)

### Statistics

We acquired descriptive statistics using cross-tabulations. Nominal values were presented as counts (percentages). Continuous values were presented as medians or means based on whether the values complied with Gaussian distribution. Medians were reported with interquartile range (IQR) for large groups or range for smaller (n < 50) groups. Means were reported with standard deviation (S.D.). The normality of continuous values was assessed visually using histograms and Q-Q plots and with skewness value of the distribution. For the statistical analysis, we used the statistical program SPSS 28.0.1 (IBM corp. released 10 November 2021).

### Ethics and approval

An organizational approval was sought for all participating hospitals. An ethical approval was not required as the study was retrospective and did not require interaction with the patients.

## Results

We identified 466 e-scooter-related injuries, of which 446 fulfilled our inclusion criteria (Figure 1). In 434 cases, the injured person was the rider, whereas in 12 cases, a pedestrian or a cyclist was injured. The median age of the patients was 26 years (IQR 22-33, range 16-92) for riders and 46 years (range 17-66 years) for non-riders. 260 (59 %) riders injured were male (Table 1). In 67 cases, the e-scooter involved in the accident was reported to be a shared e-scooter, in 17 cases the e-scooter was privately owned, and in 362 cases the information regarding e-scooter ownership was not available.

**Table 1:** Patient and injury characteristics of the e-scooter injuries. The reported values are counts (%) on binominal variables and medians (IQR) for continuous variables, if not otherwise specified.

|  | **Riders**<br>(n = 434) | **Non-riders**<br>(n = 12) | **Total**<br>(n =446) |
| --- | --- | --- | --- |
| **Age, years, median (range)** | 25.9 (16.2–91.8) | 46.1 (17.0–65.5) | 26.0 (21.6–32.8) |
| **Male sex** | 255 (58.8 %) | 5 | 260 (58.3 %) |
| **Alcohol intoxication** | 198 (45.6 %) | 3 | 201 (45.1 %) |
| Breath alcohol, mean (S.D)<br>(n=162) | 1.5 (0.7) | N/A | 1.5 (0.7) |
| **Helmet usage** | 14 (3.2 %) | N/A | 14 (3.1 %) |
| **AIS<sup>a</sup>** |  |  |  |
| Minor | 247 (56.9 %) | 10 | 257 (57.6 %) |
| Moderate | 153 (35.3 %) | 2 | 155 (34.8 %) |
| Serious | 30 (6.9 %) | 0 | 30 (6.7 %) |
| Severe | 3 (0.7 %) | 0 | 3 (0.7 %) |
| Critical | 1 (0.2 %) | 0 | 1 (0.2 %) |
| **ISS<sup>b</sup>** |  |  |  |
| Minor (1-8) | 400 (92.2 %) | 12 | 412 (92.4 %) |
| Moderate (9-14) | 30 (6.9 %) | 0 | 30 (6.7 %) |
| Major (>15) | 4 (0.9 %) | 0 | 4 (1.1 %) |
| **The use of EMS<sup>c</sup>** | 185 (42.6 %) | 4 | 189 (43.3 %) |
| **Initial place of treatment** |  |  |  |
| Level I trauma center | 82 (18.9 %) | 2 | 84 (18.8 %) |
| Level IV trauma center | 352 (81.1 %) | 10 | 362 (81.2 %) |
| **ICU<sup>d</sup> admission, n (%)** | 5 (1.2 %) | 0 | 5 (1.1 %) |
| Length of stay, days, median<br>(range) | 2 (1–8) | N/A | 2 (1–8) |
| **Hospital ward admission, n (%)** | 58 (13.4 %) | 2 | 60 (13.5 %) |
| Length of stay, days, median<br>(range) | 2.5 (1–164) | 2 (1–3) | 3 (1–164) |
| **Operative treatment, n (%)** | 53 (12.2 %) | 1 | 54 (12.1 %) |

- a. Classification of injuries by Abbreviated Injury Score - b. Classification of injuries by Injury Severity Score - c. EMS = Emergency medical services - d. ICU= Intensive care unit

### Injury characteristics

The most common injury mechanism was a fall (n= 374), followed by collision (n = 40). Furthermore, in 15 accidents there was two people reported on the same scooter. 13 scooter riders sustained a collision with a moving car. The non-riders were either pedestrians (n = 9) or bicyclists (n = 3) that got hit by or collided with an e-scooter rider (Table 2).

**Table 2:** Injury mechanisms. In total, there were 446 persons injured in an e-scooter-related accident

|  | N (%) |
| --- | --- |
| **Rider injured** | 434 (97.3 %) |
| The rider fell | 374 (83.9 %) |
| Non spesified | 353 (79.1 %) |
| Tripped over a curb | 18 (4.0 %) |
| Two persons on a same scooter fell | 15 (3.4 %) |
| Tripped over a tram rail | 3 (0.7 %) |
| Collision | 40 (9.0 %) |
| with an object | 19 (4.3 %) |
| with a moving car | 13 (2.9 %) |
| with a cyclist | 4 (0.9 %) |
| with another e-scooter | 4 (0.9 %) |
| Injured while riding<sup>a</sup> | 5 (1.1 %) |
| **Non-rider injured** | 12 (2.7 %) |
| Pedestrian hit by e-scooter | 9 (2.0 %) |
| Bicyclist collided with an e-scooter | 3 (0.7 %) |

According to abbreviated injury score 58 % (n = 257) of injuries were considered minor, whereas 35 % (n = 155) were moderate, and 8 % (n = 34) serious, or worse. Three patients had severe injuries, of which two were intracranial hemorrhages, and one was internal bleeding requiring angioembolization. One patient sustained a critical traumatic brain injury requiring an emergency craniectomy. The non-riders sustained only minor (n = 10) or moderate (n = 2) injuries. Distinct categorizations of the injury severity are presented in table 1.

The most common injury site was head and face with 220 (49 %) injured patients. Of these patients, 50 (11%) were diagnosed with a concussion, and 16 (3.6 %) sustained an intracranial hemorrhage. The upper and lower extremities were injured in 142 (32 %) and 113 (25 %) patients, respectively. Excluding minor injuries, the most common injuries were fractures of the hand (n=31) and distal forearm (n=13). Including all locations, 151 (34 %) patients sustained fractures. The injuries are presented in detail in figure 2.

**Figure 2:**
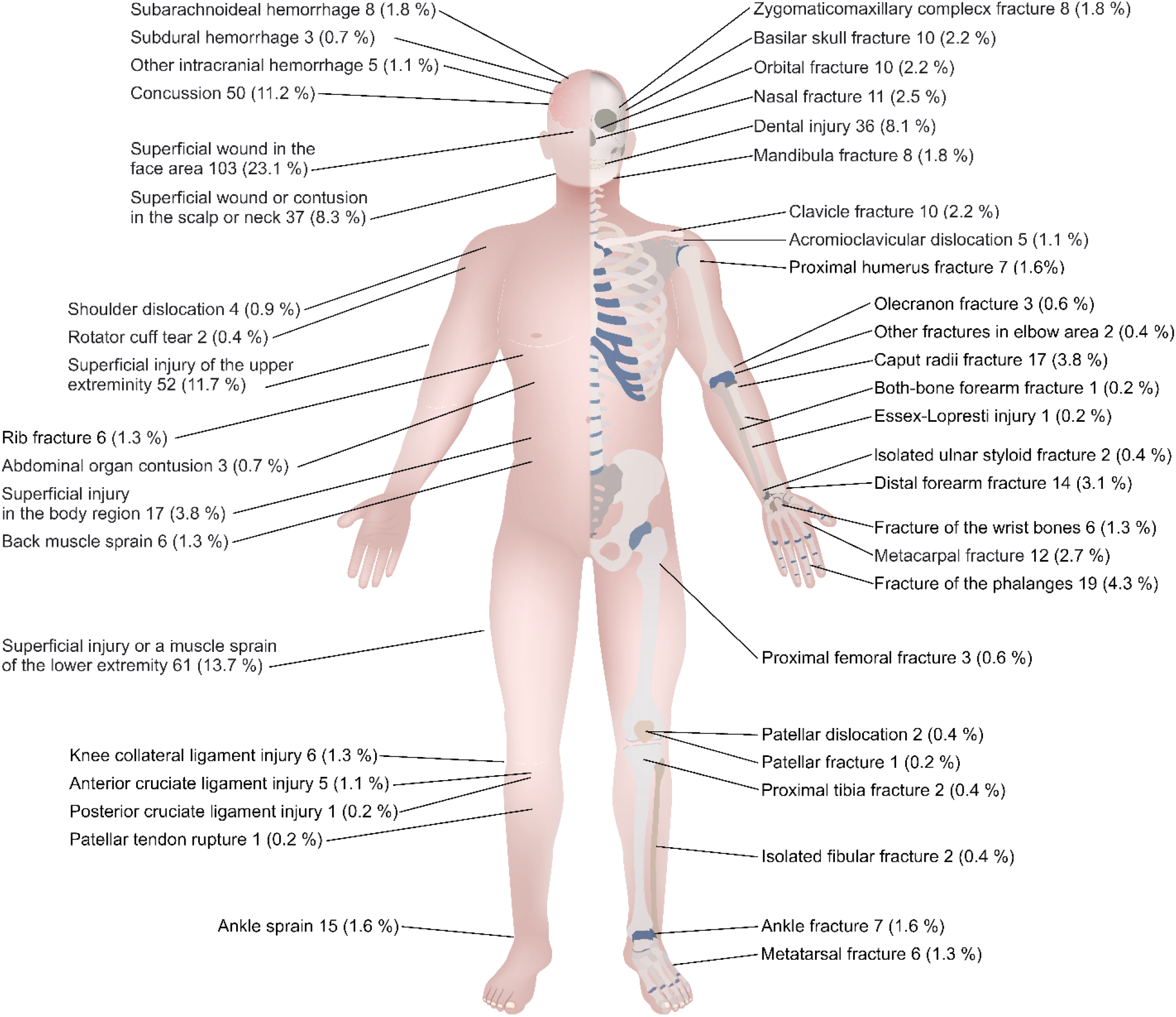
Anatomical locations of the injuries sustained in e-scooter accidents.

### Affecting factors and temporal distribution

In total, 201 (45 %) patients were reported to be intoxicated by alcohol at the time of the injury. The effect of intoxication was emphasized during night-time as 75 % of the patients injured between 00.00 and 5.00 were reported to be intoxicated (Figure 3). Helmet use was reported in 14 (3 %) riders, although in 303 (70 %) cases, there was no definite information available on helmet use.

**Figure 3:**
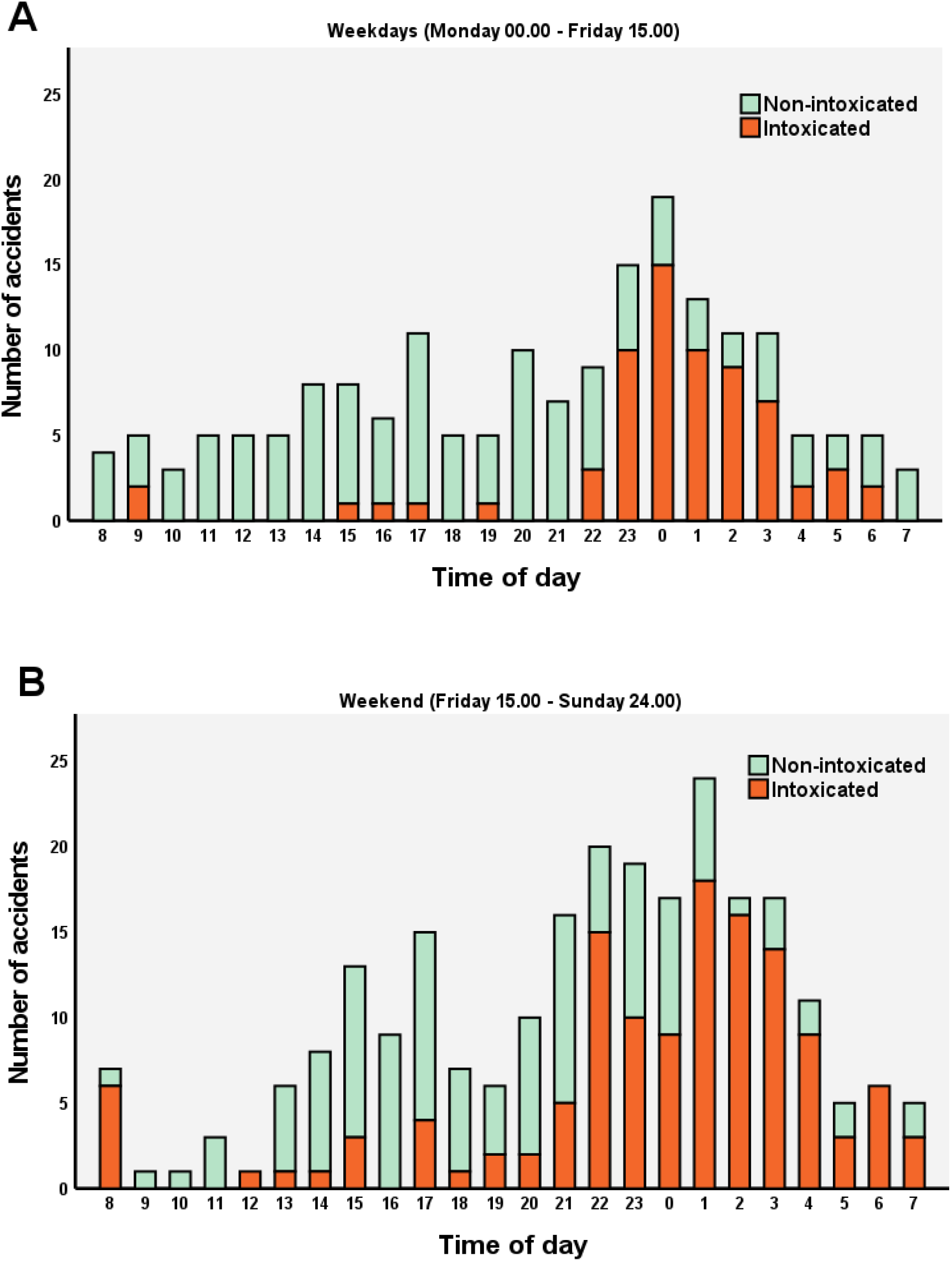
Histogram of time of day of the accidents and proportion of alcohol intoxication. 5A) Accidents during weekdays (From Monday 00.00 to Friday 15.00) 5A) Accidents during weekends (From Friday 15.00 to Sunday 24.00)

Most of the accidents happened in July (n = 131), followed by June (n= 101) and August (n = 75), respectively (Figure 4). 51 % (n = 227) of the accidents happened before the speed restrictions on the inner-city, and 84 % (n = 375) before weekend night-time ban, respectively. In total, 55 % (n = 246) of the accidents happened during the weekend (Friday 15:00 to Sunday 24:00) (Figure 5). There was a major spike in accident incidence during weekend late evenings, and nights (Figure 3).

**Figure 4:**
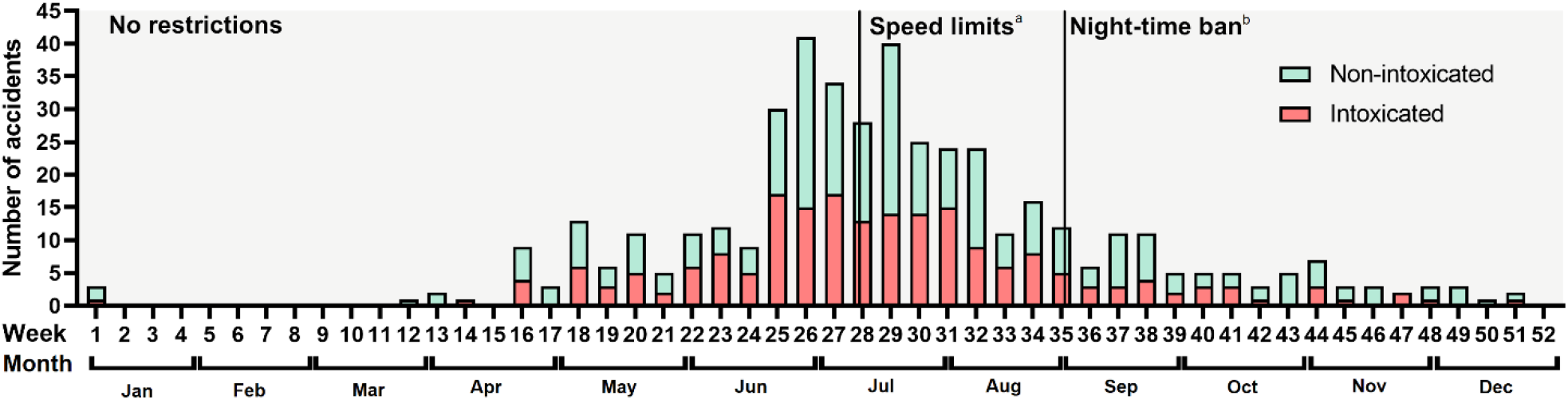
E-scooter accidents during the year reported by weekly accident incidence. The restrictions in operation are shown. a. Speed restrictions of 15 km/h in certain the inner-city locations b. Shared e-scooter services rental ban on Fridays and Saturdays between 00.00 and 05.00. In addition, the top speed on was set to 20 km/h during daytime and to 15 km/h between 00.00 and 05.00 (except on Fridays and Sundays)

**Figure 5:**
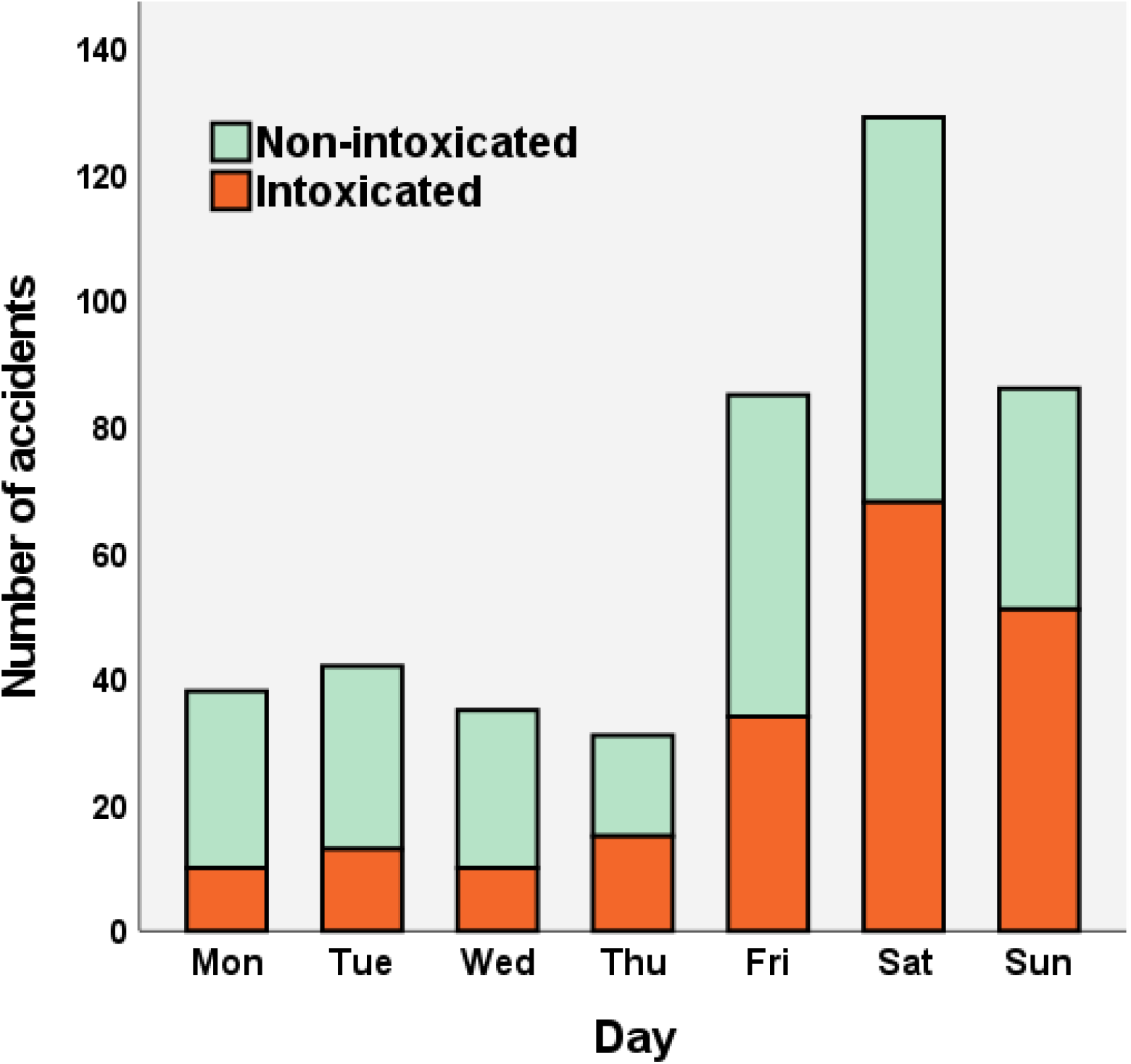
Histogram of accidents during the week and proportion of alcohol intoxication

### Aftercare

Overall, 60 (13 %) patients were admitted to a hospital after their ED visit, 2 of which were non-riders. 5 riders required intensive care, with median length of ICU treatment being 2 days (range 1-8). The rest were admitted directly to traditional hospital wards. The median length of stay in the hospital was 2 days (IQR 1-5 days).

Overall, 189 (42 %) patients required additional doctor appointments after their primary ED visit, whereas 309 patients (69 %) visited some type of healthcare professional (including doctors’ appointments physiotherapy, nurse appointments, dental appointments and psychologist appointments)

Operative treatment was required in 53 (12 %) patients. In total, 60 operations were made in the follow-up period. Orthopaedic procedures on extremities were most common (n = 46), with operations on distal forearm fractures (n = 7), fractures of hand bones (n = 6), and ankle fractures (n = 5) being the most frequent.

In addition, 8 maxillofacial and 5 neurosurgical procedures were performed. A single patient required a radiological angioembolization due to major bleeding in the splenic artery (Table 3).

**Table 3:** Operative treatment of the injuries. In total, there were 60 operations on 53 patients.

|  | N |
| --- | --- |
| **Orthopaedic operations** | 46 |
| Upper extremity operations | 26 |
| Distal forearm fracture fixation | 7 |
| Hand bone fixation | 6 |
| UCL stabilization in MCP 1 | 3 |
| Proximal humerus fracture fixation | 2 |
| Shoulder ligament stabilization | 2 |
| Clavicle fracture fixation | 2 |
| Other operations on upper extremities<sup>a</sup> | 4 |
| Lower extremity operations | 20 |
| Ankle fracture fixation | 5 |
| Proximal tibia fixation | 4 |
| Proximal femoral fracture fixation | 3 |
| Knee ligament reconstruction via arthroscopy | 3 |
| Other operation on lower extremities<sup>b</sup> | 5 |
| **Maxillofacial operations** | 8 |
| Zygomaticomaxillary complex fracture fixation | 3 |
| Mandibula fracture stabilization | 4 |
| Le Fort Maxilla fracture stabilization | 1 |
| **Neurosurgical operations** | 5 |
| Craniectomy | 2 |
| Other neurosurgical operations<sup>c</sup> | 3 |
| **Other operations** | 1 |
| Angioembolization of splenic artery | 1 |

a. Diaphyseal both-bone fracture fixation, Essex-Lopresti stabilization, Olecranon fracture fixation, Plate removal
b. Patellar tendon repair, Tibia diaphyseal fracture fixation with a cephalomedullary nail, Surgical wound revision (n=3)
c. Cranioplasty, Neurosurgical wound revision surgery

### Cost analysis

The total cost of hospital care, including ED visits, imaging, inpatient care, and surgeries, and follow-up visits was approximately 866 889 €. A median of 1059 € (IQR 296 – 1966 €) was billed per patient.

Based on the physician’s statements, a total of 13.5 years (4928 days) worth of sick leave was prescribed for 178 patients. Of those affected, the median length of sick leave was 14 days (IQR 5-38 days). In total, this inflicted an estimated 284 047 € of expense for the work providers and 561 464 € for the Social Insurance Institution of Finland.

Including the cost of the hospital care and follow-up as well as the prescribed sick leaves, the cumulative cost of the e-scooter injuries was approximately 1.71 million euros, with a median cost of 1148 € (IQR 399 – 4263 €) per patient. A detailed presentation of the economic impact of e-scooters is found in Table 4.

**Table 4:**
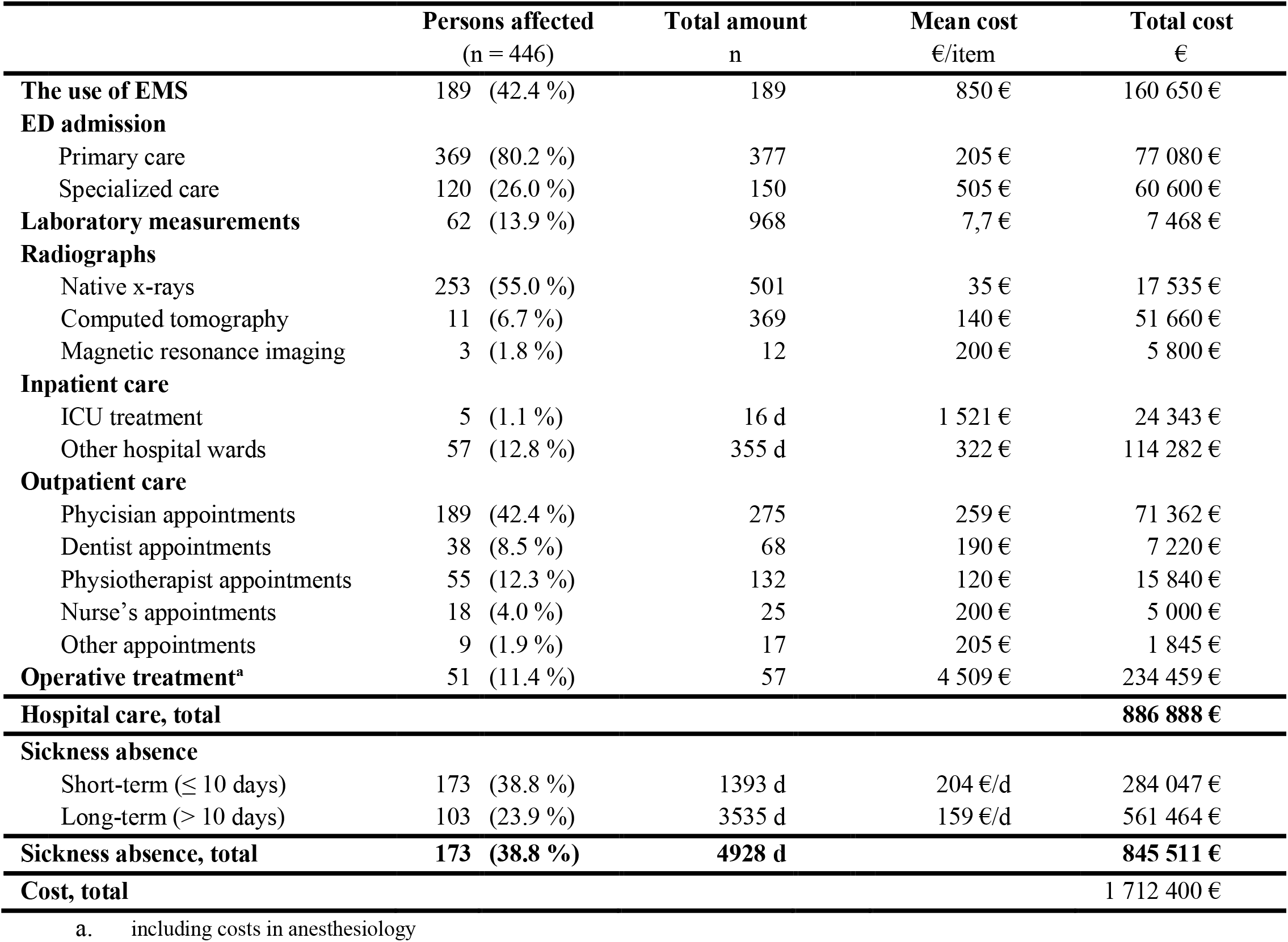
Cost analysis for e-scooter injuries

## Discussion

Although e-scooters are a convenient emerging mode of transport, the risk for injuries enhances the disadvantages. This study has demonstrated that while most e-scooter injuries are minor, 42 % of the injuries are moderate, severe, or worse. 13 % of the patients in our material required hospital admission, and 11 % required surgical interventions. Furthermore, the median cost of an injury was 1148 € with a total cost of 1,7 million €. This research provides insights for the decision-making on whether the price and loss in health are worth the usefulness of a new emerging travel mode.

Our findings on injury characteristics complement those of earlier publications. It is well established that people presenting themselves to the ED with an e-scooter injury are most often males (54-67 %) in their late twenties or early thirties (Bekhit et al., 2020; Bloom et al., 2021; Coelho et al., 2021; Farley et al., 2020; Lavoie-Gagne, Siow, Harkin, Flores, Girard, et al., 2021; Moftakhar et al., 2021; Namiri et al., 2020; Shichman et al., 2022; Siow et al., 2020; Trivedi et al., 2019). In 92-96 % of the cases, the injured are the riders themselves (Coelho et al., 2021; Ingstrup Nielsen et al., 2021; Shichman et al., 2022; Trivedi et al., 2019). As was also seen in our material, the most common site of injury is typically the head (27-47 % of the cases) (Farley et al., 2020; Ingstrup Nielsen et al., 2021; Moftakhar et al., 2021; Namiri et al., 2020; Trivedi et al., 2019; Uluk et al., 2022), although in some publications the extremities have been the predominant site (Bloom et al., 2021; Coelho et al., 2021). Nevertheless, a growing body of published work indicates that e-scooter injuries’ characteristics are well known.

A notable finding of our study was that a predominant number of accidents happened during weekends and night-time. Unfortunately, a greater portion of patients was also intoxicated during these times. Similar findings have also been reported in previous publications, although the correlation between alcohol usage and weekend night-time is not well established in these publications (Blomberg et al., 2019; Moftakhar et al., 2021; Suominen et al., 2022; Uluk et al., 2022). Although driving while intoxicated is forbidden by the law and the rules of shared e-scooter companies, effective surveillance is not possible. As a result, the city of Helsinki and the e-scooter rental companies banned the use of shared e-scooters during the night-time (00.00-05.00) in the beginning of September 2022. Anderson et al. examined the effect of a similar night-time ban on e-scooter rentals (9 p.m. to 4 a.m.) in city of Atlanta (USA), after which a decrease in e-scooter accidents was seen. However, the effect on ED time of arrival was not significant in their study (p = 0.16). (Anderson et al., 2021) When taking our results into consideration, the night-time ban appears to be justifiable. However, extensive conclusions cannot be drawn as the beginning of autumn, and the cooling of the weather also decreases the e-scooter usage drastically.

Mandatory helmet use has been brought up in many publications (Blomberg et al., 2019; Coelho et al., 2021; Crowe & Elkbuli, 2021; Moftakhar et al., 2021). In our material, nearly half of the patients sustained a head injury. These varied from mild contusions to life threatening intracranial bleeding. Similar to our material, the reported helmet usage has been typically 2-4 % in the injured (Blomberg et al., 2019; Bloom et al., 2021; Coelho et al., 2021; English et al., 2020; Siow et al., 2020; Trivedi et al., 2019). In countries where helmet use is mandatory, helmets have been reported in 20-45 % of the injured (Coelho et al., 2021; Mitchell et al., 2019). On the other hand, compulsory helmet use seems to decrease the usage of e-scooters (Lo et al., 2020). From the standpoint of injury prevention, this would be desirable, although this would limit the users’ freedom. Moreover, the effectiveness of helmet usage is not well studied on e-scooters. Mitchell et al. found in their small-scale study (n = 54) that helmet usage reduced the presence of head injuries with OR of 0,18 (95 % CI 0,04-0,83) (Mitchell et al., 2019). In comparison, the preventive effectiveness of helmet usage is well established in cycling, where it is estimated to provide a 63 to 88 % reduction in head injuries (Thompson et al., 1999). Thus, we believe helmets should be strongly couraged on e-scooters to prevent severe head injuries.

The estimates for costs for e-scooter injuries vary depending on the country. For example, Bekhit et al. estimated in New Zealand, that the cost of a single e-scooter injury was approximately 1 000 € (1693 NZD) (Bekhit et al., 2020) whereas in the United States average cost is estimated to be between 1100 € (1213 USD, median) and 90 000 € (95 710 $, mean)(Bloom et al., 2021; Lavoie-Gagne, Siow, Harkin, Flores, Politzer, et al., 2021). Furthermore, in Australia, the average cost was determined as 364 € (Mitchell et al., 2019). Unquestionably, the methods for cost estimations vary, but undoubtedly, e-scooter usage comes with a cost.

While the present study was made in precision, there are also several weaknesses. First, as we did not have an exact ICD-code for e-scooter injuries, we had to inquire the patients via a keyword search. As a result, some e-scooter injuries might have gone unnoticed. Nonetheless, the number of patients was large, and the loss of patients can be considered random. Thus, a selection bias can be regarded as minimal, and the injury proportions are accurate. Second, due to the retrospective nature of the data, the distinction between vehicles could not be made completely accurately. There is a possibility that the ED personnel might have miscategorized some vehicles. Furthermore, the distinction between privately owned and shared e-scooters was not possible. As there are no distinct protocols, the amount of helmet usage and alcohol intoxication might be underreported. Third, some milder injuries are also treated in private hospitals. Thus, the reported incidence underestimates the actual hospital requiring e-scooter injury rate. However, the hospitals in the study treat the vast majority of ED requiring accidents in our city, and therefore the reported incidences are solid estimates of the actual incidence.

The main strength of the study is that while many studies focus on a restricted pool of patients (e.g., patients of a single hospital, craniofacial injuries, or injuries requiring surgical care), we report all emergency care requiring e-scooter injuries from the whole city – varying from mild trauma to life-threatening accidents. In addition, we performed a comprehensive estimation of the economic impacts of the e-scooters. While there are studies with a greater quantity of patients, the studies exploit national registries with a larger number of inaccuracies. (Farley et al., 2020). All things considered, the present study adds considerably to the current knowledge of e-scooter-related injuries.

## Conclusions

While most e-scooter injuries are minor, a considerable proportion of the injuries are moderate, severe, or worse, including patients requiring intensive care and operative treatment. Comprehensive preventive measures must be conducted to decrease the incidence of e-scooter injuries. The night-time bans and speed-limits on rental e-scooters during weekends appear to be justifiable means to decrease the disadvantages - both in individual and social perspective.

## Data Availability

The datasets generated and analysed during the current study are not publicly available due potential other publications utilizing the data but are available from the corresponding author on reasonable request.

## List of abbreviations

E-scooter: Stand up electric scooter
ED: Emergency department
AIS: Abbreviated Injury Score
ISS: Injury Severity Score
IQR: Inter Quartile Range
EMS: Emergency Medical Services
ICU: Intensive Care Unit

## Declarations

Not applicable

## Ethics approval and consent to participate

An organizational approval was sought to access the data from all participating hospitals (HUS/44/2021). Ethical committee was not consulted as the study was retrospective and did not require any interaction with the patients.

## Consent for publication

Not applicable

## Authors’ contributions

HV conceptualized and planned the core study design. VH, KV, MC, and AK reviewed and proposed improvements for the study design. HV and LT performed the data enquiry. HV analyzed the data, performed the statistical analysis, and wrote the core manuscript. All authors contributed to the interpretation of the data and manuscript revision and approved the final manuscript.

## Acknowledgments

The authors wish to thank the E-scooters in Helsinki -working committee, including the City of Helsinki, Ministry of Transport and Communications, and the e-scooter rental companies, led by Milos Mladenovic, for their active surveillance and proceedings regarding the e-scooter accidents. In addition, we thank Sini Hilve for the help with Figure 2.

## Authors’ information

HV: Junior ED doctor, PhD student

LT: Bachelor of Medicine

VH: Consultant in Emergency Medicine, Cardiology and Internal Medicine, Head Physician of Emergency Medicine and Service, PhD, the title of docent

KV: Consultant in Orthopaedics and Traumatology, PhD

MC: Consultant in Anesthesiology and Emergency medicine, Director of Emergency Medicine and Services, Professor of Emergency Medicine, PhD, the title of docent

AK: Specialist in General Practice, Head of the department in basic level Emergency services

